# EEG to fMRI Synthesis for Medical Decision Support: A Case Study on Schizophrenia Diagnosis

**DOI:** 10.1101/2023.08.07.23293748

**Authors:** David Calhas, Rui Henriques

## Abstract

Electroencephalography (EEG) measures the neuronal activity at the scalp, while functional magnetic resonance imaging (fMRI) provides a sub-cortical view of blood supply in the human brain. Although fMRI is known for providing rich spatial information, it is expensive and of restricted use. EEG to fMRI synthesis is a cross modal research area that bridges the gap between the two and has recently received attention. Although these studies promise lower healthcare costs and ambulatory assessments, their utility in diagnostic settings is still largely untapped. Using simultaneous EEG and fMRI recordings, this study combines a state-of-the-art synthesis model with a modified contrastive loss, and subsequent prediction layering, to unprecedentedly assess its predictive power in schizophrenia diagnosis. In addition, we perform an exhaustive search for the (synthesized) hemodynamic brain patterns able to discriminate schizophrenia. Schizophrenia diagnosis using synthesized hemodynamics yield an area under the ROC curve of 0.77, confirming the validity of the undertaken neuroimaging synthesis. Experiments further revealed schizophrenia-related patterns in frontal, left temporal and cerebellum regions of the brain. Altogether, our results suggest that a synthesized fMRI view is able to discriminate this pathology, and it contains discriminative patterns of brain activity in accordance with related work on schizophrenia.

## 1 Introduction

Schizophrenia is a neurological condition that affects a significant portion of the world population [1]. Yet, to this day, the diagnosis of schizophrenia generally involves several clinical tests, making it time consuming and exhausting for the patient. Diagnostic tests range from psychological symptoms to neuroimaging [2], molecular [3], and natural speech markers [4]. In particular, functional magnetic resonance imaging (fMRI) scans have shown potential for an automated diagnostic [5, 6, 7, 2], still their availability is limited. For instance, Ogbole et al. [8] report the density of MRI machines in Africa, claiming Nigeria to be the most critical country, where 0.3 MRI machines are made available for each one million people. As a consequence, the quality of these health care systems is low. Can alternative cheaper modalities be considered as a proxy to replace MRI machines? Recent advances in cross modal synthesis [9, 10, 11] show promise. And although, these do not enable the replacement for cost reductions, there are claims that electroencephalography (EEG) has predictive power over fMRI features [12, 13]. Opportunely, *EEG to fMRI synthesis* is an emerging research area that focuses on the synthesis of fMRI, an expensive modality, from EEG, a lower cost modality [14, 15, 16, 17]. In spite of these great efforts, we are yet to see their applicability in an health care setting.

The main idea of this paper is to show the ability to classify schizophrenia using a synthesized fMRI from EEG, ultimately enabling better interpretation, while ensuring competitive predictive accuracy. By doing this, we provide a prediction in a feature space with fine spatial resolution. In addition, to assess its fidelity, we compare the predictive value of fMRI estimates for the target schizophrenia classification task against (state-of-the-art) time-based and frequency-based EEG views. A view is defined as a set of related features with EEG as a source (see Section 1.2). In addition to the classification problem, the synthesized fMRI is subject to a data mining tool, known to extract biclusters [18]. These biclusters identify which areas of the brain are associated with schizophrenia. In a sense, EEG, which is recorded at the scalp level, is projected to an fMRI, where sub-cortical activity patterns are retrieved. Therefore, EEG to fMRI Synthesis enriches the EEG modality with fMRI learned features [17]. This additional information increases interpretability, which we measure through a pattern discovery algorithm. The contributions of this study are:

- integration of a state-of-the-art EEG to fMRI synthesis model [17] (see Section 1.2.3) in a schizophrenia classification setting (see Section 2);
- combination of a modified contrastive loss along with layer normalization, that together allow separation of data and maintain fMRI volume style (see Section 2.1);
- biclustering analysis of a synthesized modality, which allows the retrieval of statistical significant patterns for schizophrenic individuals (see Section 3.3).

We list the main findings of this work as:

- EEG frequency features are highly discriminative of schizophrenia, which goes in accordance with previous studies [19, 20]. Nonetheless, despite its predictive power, it lacks in interpretability. Restricting its use for medical diagnostics; the decision making of the linear classification is correlated with synthesized frontal lobe activity. It is known that the functions, related to this region, are impaired by schizophrenia [21, 22]. This suggests, that the classifier founded its predictions on frontal activity;
- the cerebellum region of the synthesized fMRI is present in several biclusters associated with schizophrenia. This finding goes in accordance with MRI studies on this pathology [23];
- schizophrenia diagnosis from the synthesized fMRI, in the absence of any other complementary clinical records, is able to reach the 0.77 AUC mark, offering preliminary evidence of the validity of the cross-modal synthesis process.

### 1.1 Problem description

Let 𝒱 = {*v*_1_, …, *v*_*n*_} be a set of different views, such that ∀*i* ∈ {1, …, *n*} : *v*_*i*_ = {*F*_*i*_, *θ*_*i*_}. A view *v*_*i*_ is characterized by a function structure, *F*_*i*_, and its parameters, *θ*_*i*_. Each projection 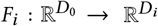 is performed from the original feature space,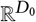, to its view space, 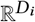. The original view is defined as 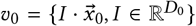, being *I* the identity matrix. Each view has a dedicated section (1.2.1, 1.2.2 and 1.2.3), where the structure of the view space, *D*_*i*_, is defined. Each instance, 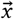, is paired with a label *y* ∈ {0, 1}^*K*^ : ∑_*k*_ *y*_*k*_ = 1, where *K* is the number of classes described in the data. A view, *v*_*i*_, optimizes its parameters, *θ*_*i*_, in order to minimize

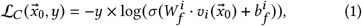

where 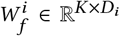 and 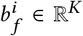 are the parameters of a linear classifier, referred to as 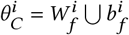, using the softmax activation, *σ*. Then the objective, for each view *i*, is defined as

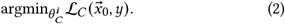

### 1.2 Background: Defining the EEG views

This study is concerned with providing an accurate view that allows a straightforward interpretation. The interpretation of a prediction is typically made by a human being, with knowledge in the application domain [24]. However, when a prediction is made by a model, it should be supported with an explanation consisting on a subset of the features 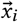 that justify the decision of the model. In this section, we describe each EEG view that is considered in our experiments. Starting with the raw (channel-temporal) view, *raw*, moving to the frequency (channel-time-frequency) representation, referred to as *stft*, and finishing with the synthesized fMRI which is the *fmri* view. For each of these, we discuss their ability to provide good explanations, as well as their overall fit to discriminate schizophrenia.

#### 1.2.1 raw view

In its raw form, an EEG recording consists of a set of electrodes, also called channels, that contain the electrical activity present at the scalp. This activity has its source at the neurons, where the action potentials occur. Formally this view is defined as *v*_0_ = {*F*_0_, *θ*_0_} where 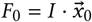 and 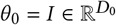.

With EEG being a multivariate time series representation, researcher,s are able to study functional properties of the brain [25]. However, the application of this specific view is mainly useful for epilepsy detection [26]. Structurally, this representation is defined as 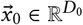, with *D*_0_ = *C* × *T*. *C* stands for the number of channels and *T* defines the temporal dimension. It is worth noting that this view is limited to the channel-temporal features, characterized by noise and difficult medical interpretation [27]. In consequence, it is unsuitable for diagnosis. In the scope of schizophrenia classification, even when processed by models with a high level of feature engineering [28], it falls short from other more fit views [29].

#### 1.2.2 stft view

Research studies, along the years, have had a common trend of extracting time frequency features from the EEG signal. From functional connectivity relations [30, 31, 32] to statistical significance of bands associated with pathologies [20, 33], several studies have used the frequency domain to uncover new findings. A time frequency representation of EEG can be achieved via a transform [34]. In this study we consider the short time Fourier transform (STFT) [35], whose kernel consists on sine and cosine waves with different shifts and frequencies. As such this view is defined as *v*_1_ = {*F*_1_, *θ*_1_}, where *F*_1_ is the sum of sinusoids and *θ*_1_ are the sinusoids themselves. Similarly to *raw*, this view is a multivariate time series representation, with the channel, *C*, and temporal, *T*, dimensions belonging to its structure. On top of that, each frequency, with the dimension *F*, is represented at different time steps and channels. Hence, the function consists of 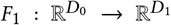 and the *stft* view is structurally defined as 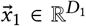, where *D*_1_ = *C* × *F* × *T*. On the interpretation side, we consider it not just similar to the *raw* view, but also more limited due to the requirement of frequency domain knowledge. Though, its discriminative ability is one of the best for schizophrenia [29].

#### 1.2.3 EEG to fMRI synthesis: fmri view

We now describe the EEG to fMRI synthesis model proposed by Calhas et al. [17]. It is a neural network, whose flow is illustrated in Figure 1. Its input is the EEG time-frequency representation view, 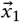. On the opposite side, the output is the *fmri* view, 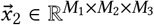, characterized by the referential axes dimensions (*M*_1_, *M*_2_ and *M*_3_). Zooming out of the neural network, we can describe this view as *v*_2_ = {*F*_2_, *θ*_2_}. Where *F*_2_ = *F*_1_ ⋃ *F* denotes the union of two functions *F*_1_, the short time Fourier transform view *v*_1_ = {*F*_1_, *θ*_1_}, and *F*, the neural network. Accordingly, its parameters are *θ*_2_ = *θ*_1_⋃ *θ*_*F*_, where *θ*_*F*_ are the parameters of the neural network.

**Figure 1.**
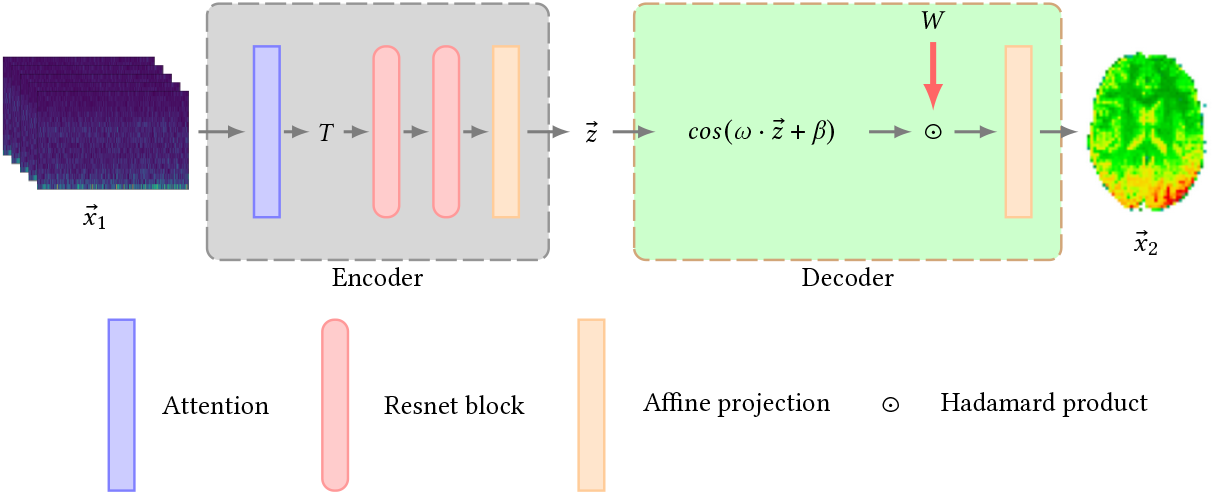
The neural architecture has two components: an Encoder (shaded in grey) and a Decoder (shaded in green). The input is the *stft v*_1_ representation. The output is the synthesized *fmri*. The Encoder begins with a simple attention mechanism on the channels dimension of the *stft*. After, it is processed by two Resnet blocks and an affine layer. This produces the latent representation 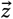. Following, comes the Decoder, which picks this representation and builds the cosine bases through the projection 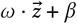. The sinusoids are style induced with 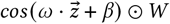. The vector *W* is a style fixed pretrained vector of an *fmri* representation, learned from a simultaneous EEG and fMRI dataset (described in Section 3.1). Finally, an affine layer projects it to the *fmri* space.

In terms of structure, this network processes the input with an attention mechanism. For this, an attention weight matrix *A* ∈ ℝ^*C*×*F* ×*T*^ is used to compute *E* ∈ ℝ^*C*×*C*^, where ∀*i* ∈ {1, …, *C*} : 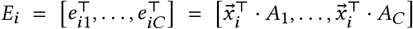. Then *E* is softmax normalized, such that

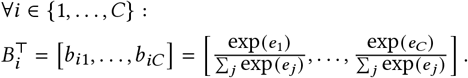

Then we can compute the attention processed representation as

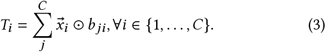

Following, *T* is processed by a set of 2 Resnet-18 blocks [36], with kernel and stride sizes tuned by a neural architecture search algorithm [37]. ^1^ These blocks contain a collection of kernels represented by *θ*_*k*_. Then, the representation is processed by a fully connected layer. All the parameters until this last layer are referred to as *θ*_*E*_, which parameterize the encoder of the neural network.

After *T* is processed by the encoder, it is then used to project random Fourier features [38, 39], parameterized by *ω* and *β*. Before decoding, the sinudoids are multiplied with a learned latent style vector, *W* ∈ ℝ^*L*^, as 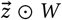. The result is processed by an affine transformation, mapping it to the fMRI volume space 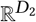. With this, we formulate the function structure *F* : 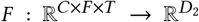. Regarding interpretability, no previous studies have assessed the quality of EEG to fMRI synthesis in decision making settings, such as schizophrenia classification. Therefore, no assumptions can be made at this stage. We try to answer this question in the end of this manuscript.

## 2 Creating discriminative fMRI views of EEG

To assess which view is better, we need to choose a classifier without a high feature engineering ability, in order to truly see the discriminative power of the view. A linear classifier fits this requirement, because it bases its decision solely on the multiplication of features with coefficients (weights), without additional feature rearrangement. To further motivate its use, a linear classifier has been used as a metric to assess explanation quality [24], exactly due to its lack of feature engineering ability.

As such, a linear classifier is used for the considered views. But a special case for *v*_2_ arises. Since this view is produced by sinusoids, it disables the separation of data according to the input 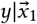. We consider that, for generalization sake, the EEG modality contains information to separate the data, 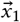, according to its labels, *y*. Though, if this representation has its distribution broken at the sinusoids, we can no longer use it. To avoid this, we propose a method that maintains the sinusoids in the neural architecture flow and manipulates the representation, so that the separation before the sinusoid, 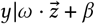, is identically distributed to 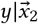.

### 2.1 Sinusoid separation

In order to separate data along the projection of cosines, we have to operate in a sub-domain interval where the *cos* is not periodic. Layer normalization [40], with center in 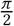 and standard deviation of 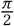, maps most of the data to such an interval of the cosine. See Figure 2 for an illustration example.

**Figure 2.**
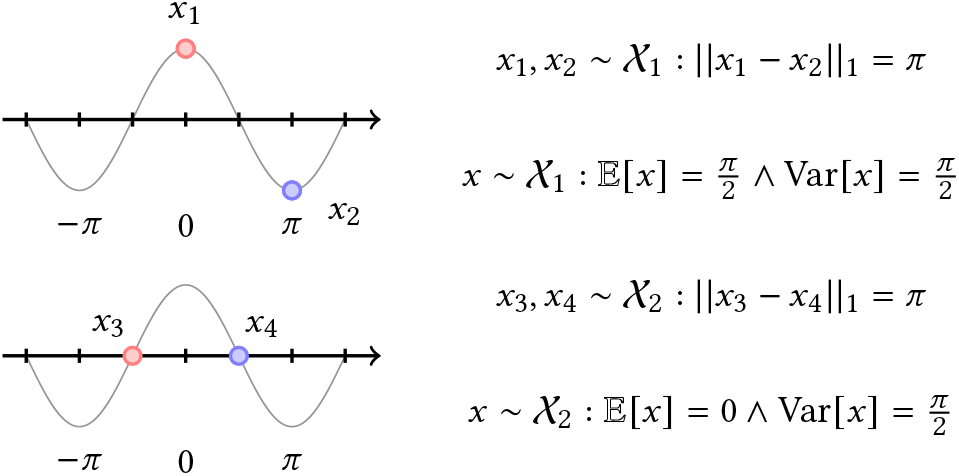
Description of how two similarly distributed samples, taken from 𝒳_1_ and 𝒳_2_, can lead to different portions of the cosine function image, since a sinusoid is periodic. Two distributions 𝒳_1_ and 𝒳_2_, may be mapped to the same image (second/bottom example). This is why a cosine is a shift invariant function. However, there are intervals a shift can be made and it is not invariant. Such intervals take the form ∀*i* ∈ ℤ : [*iπ*, (*i* + 1)*π*].

Nonetheless, the assurance that the projections are likely in a non periodic sub-domain of the cosine function, does not alone complete separation of the data. This is because the distribution 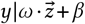 needs to be separated accordingly. We propose a modified contrastive loss defined as

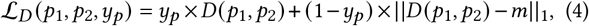

where *p*_1_, *p*_2_ formulate a pair of two instances and *y*_*p*_ ∈ {0, 1} defines the pairwise label, being 0 when the label of *p*_1_, *p*_2_ mismatch and 1 when they match. *D* (*p*_1_, *p*_2_) is the *l*1 distance 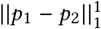. Setting *m* ∈ [*ϵ, π*] : *ϵ >* 0, together with the described layer normalization, allows separation of the data within a non periodic sub domain of the cosine. In Figure 3, the effects of this methodology are illustrated, before and after the learning session.

**Figure 3.**
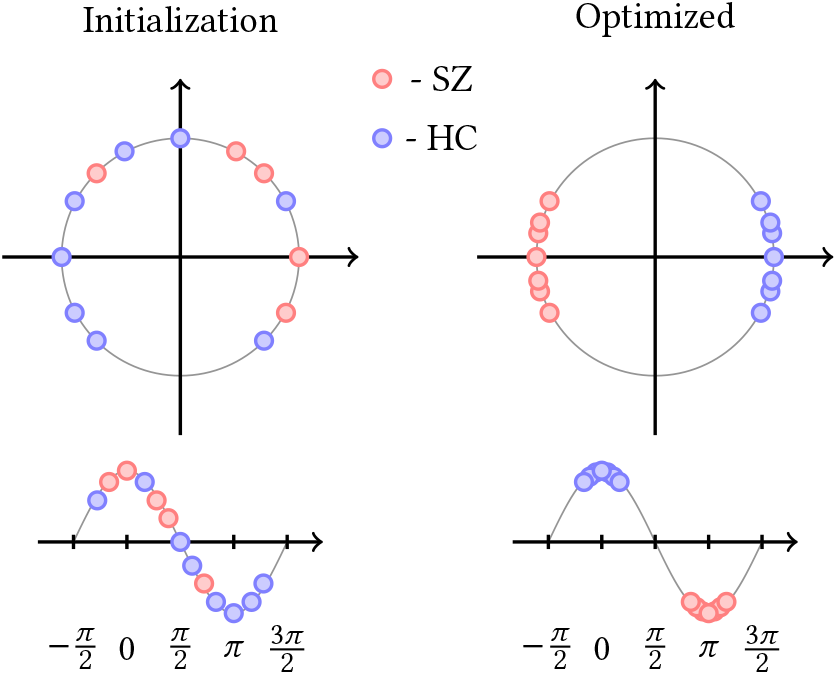
Normalization of data points inside the unit circle, using layer normalization, along with the optimization of a contrastive loss lead to correct separation of sinusoids. Data points belong to two classes, *HC* and *SZ*, that are separated after the minimization of ℒ_*D*_. Because we separate false pairs, according to (1 − *y*_*p*_) × ||*D*(*x*_1_, *x*_2_) − *m*||_1_, all points are placed within a shift variant interval of the cosine. The variance needed for classification.

## 3 Experimental setting

In our experiments we considered 3 EEG views: 1) natural/raw EEG view in ℝ^*C*×*T*^, referred to as *v*_0_; 2) time-frequency representation of EEG in ℝ^*C*×*F*×*T*^, referred to as *v*_1_; 3) synthesized fMRI representation in 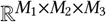, referred to as *v*_2_. The optimization of the parameters, of each of these, consists on the minimization of ℒ_*C*_, such that the gradients are 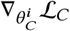. The special case *v*_2_ = {*F*_2_, *θ*_2_} requires a pretraining session in a simultaneous EEG and fMRI dataset, so it learns fMRI representations. Following, comes the minimization of the classification objective, where 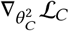 is computed along with 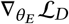, being *θ*_*E*_ the parameters of the encoder of *F* (recall Section 1.2.3). All these objectives require the respective data for the computation of the gradients described above. Namely EEG data for classification and, in particular for *v*_2_, the simultaneous EEG and fMRI data, for EEG to fMRI synthesis.

### 3.1 Datasets

#### *Classification: EEG only*, Fribourg dataset

Padée et al. [41] gathered 43 individuals, 24 healthy controls and 19 with diagnosed schizophrenia, and performed a task-based recording session. In this recording session each individual played a game. The EEG was set with 128 electrodes, distributed according to the 10-20 system. The sampling rate was 2048Hz.

#### *Synthesis: simulatenous EEG and fMRI*, NODDI dataset

Deligianni et al. [42] performed resting state recordings and a total of 8 individuals are considered. The EEG was setup with 64 channels, placed according to the 10-20 system. Recorded at a sampling frequency of 250Hz. The fMRI setup was recorded with a 2.160 Time Response and 30 millisecond echo time. Each voxel is 3 × 3 × 3 millimeters, consequently making the resolution of a volume *M*_1_ × *M*_2_ × *M*_3_ = 64 × 64 × 30 voxels.

### 3.2 Validation

To validate the hypotheses drawn, each view, *v*_*i*_, will do a validation process. To this end, the steps of this process are:

1. leave-one-individual-out cross validation (LOOCV);
2. cross validation (5 folds) hyperparameter optimization with 25 iterations for each fold of the LOOCV step;
3. particularly for fMRI synthesized views, *v*_2_, a pretraining session is done, where *F* is optimized on simultaneous EEG and fMRI data.

The hyperparameters used to train *F* are in accordance with the original work [17]. For the cross validation (step 2.), Bayesian optimization [43] is performed and the hyperparameters subject to optimization are: *l*1 regularization constant ∈ [1e − 10, 2.0], learning rate ∈ [1e − 5, 1.0] and batch size ∈ {1, 2, 4, 8, 16, 32}. The learning session is fixed with 10 epochs and gradients are propagated using the Adam optimizer [44].

### 3.3 Biclustering

With the synthesized fMRI view, 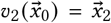, it is important to address which synthesized regions are most discriminative for schizophrenia. As well as compare the reported regions with related studies. For this, we ran BicPAMS [45] to find cluster subspaces composed of a subset of rows (individuals) and columns (voxels) that discriminate a target (pathology or healthy). Two settings are considered: 1) clustering 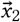features with ground truth labels *y*; and 2) clustering 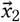 features with its linear classifier predictions 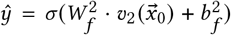. This two settings allow us to find discriminative and explainability patterns from the synthesized fMRI view.

*Discriminative* patterns provide us sets of individuals and features that support a certain target. This analysis is similar to classification, but instead of having a classifier, we have a set of association rules. In turn, *Explainability* patterns give us interpretable information about the decision made by the linear classifier. Although it does not provide discriminative patterns, it gives us association rules for the decision making process of the classifier. The biclustering of the features, 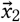, and the predictions, *ŷ*, produces global explanations, forming a novel explainability algorithm. Jointly, these settings allow us to have a better comprehension from the synthesized fMRI view, potentially able to uncover advantages and pitfalls. An important analysis when working with data driven projections. Biclusters were found in three resolutions: 5 × 5 × 3; 10 × 10 × 5; and 14 × 14 × 7. These resolutions enable us to assess patterns at different granularities. In a 5 × 5 × 3 clusters represent big regions of the brain, as big as entire lobes; a 10 × 10 × 5 resolution can still retrieve regions of interest, but at a finer granularity; and 14 × 14 × 7 goes even more detailed. Altogether, this analyses give us patterns, with statistical assurances for a target (schizophrenia).

The parameters given to BicPAMS to search for biclusters in the different resolutions are shown in Table 1. Note that, for the lowest resolution 5 × 5 × 3, the number of *columns*, which translates to the number of voxels is only 1. This is because 1 voxel in a 5 × 5 × 3 corresponds to a big region in the original resolution, 64 × 64 × 30. As the resolution increases, we also increased the number of voxels required to appear in a bicluster. The number of *voxels* pretains to the discretization of the data. It is important to not increase this parameter, as the *items-boundary* problem can arise [46]. This problem manifests when the number of *bins* is high and similar values are put in different bins. The number *biclusters* requires that all biclusters obtained are bigger than this value. The *lift* tells us how discriminative is the cluster for a target. In other words, the *lift* is the ratio of how much an item occurs in the bicluster by how much it occurs in the dataset. Typically, *lift* ≫1 indicates that the selected bicluster is able to discriminate the given target.

**Table 1.** Parameters for the biclustering algorithm.

| Parameter | $5 \times 5 \times 3$ | | $10 \times 10 \times 5$ | | $14 \times 14 \times 7$ | |
| --- | --- | --- | --- | --- | --- | --- |
| | $y$ | $\hat{y}$ | $y$ | $\hat{y}$ | $y$ | $\hat{y}$ |
| lift | 1.3 | 1.3 | 1.3 | 1.2 | 1.3 | 1.2 |
| # biclusters | 100 | 100 | 100 | 100 | 100 | 100 |
| # bins | 3 | 3 | 5 | 5 | 5 | 5 |
| min # voxels | 1 | 1 | 10 | 10 | 15 | 15 |

## 4 Results

The results for the classification experiment are shown in Table 2. The *stft* representation had the best performance with an AUC of 0.933, followed by *fmri* with 0.765 and *raw* performed below random with 0.225. The performance of the *raw* representation is low and shows that an EEG recording without preprocessing steps is not able to be applied in schizophrenia classification settings. On the other hand, the *stft* is capable of it, showing the time-frequency domain features are discriminative of schizophrenia. Nonetheless, we expected *fmri* to be closer to the performance of *stft*. An AUC of 0.765 shows it has a good prediction power, however, it was outperformed by its preceding representation, according to Figure 4. We found no statistical significance between *raw* and *fmri*, on the other hand *stft* outperformed *raw* and *fmri* with statistical significance.

**Table 2.** AUC of a linear classifier with different views as input.

| View | Fribourg (AUC) |
| --- | --- |
| *raw* $v_0$ | 0.225 |
| *stft* $v_1$ | **0.933** |
| *fmri* $v_2$ | 0.765 |

**Figure 4.**
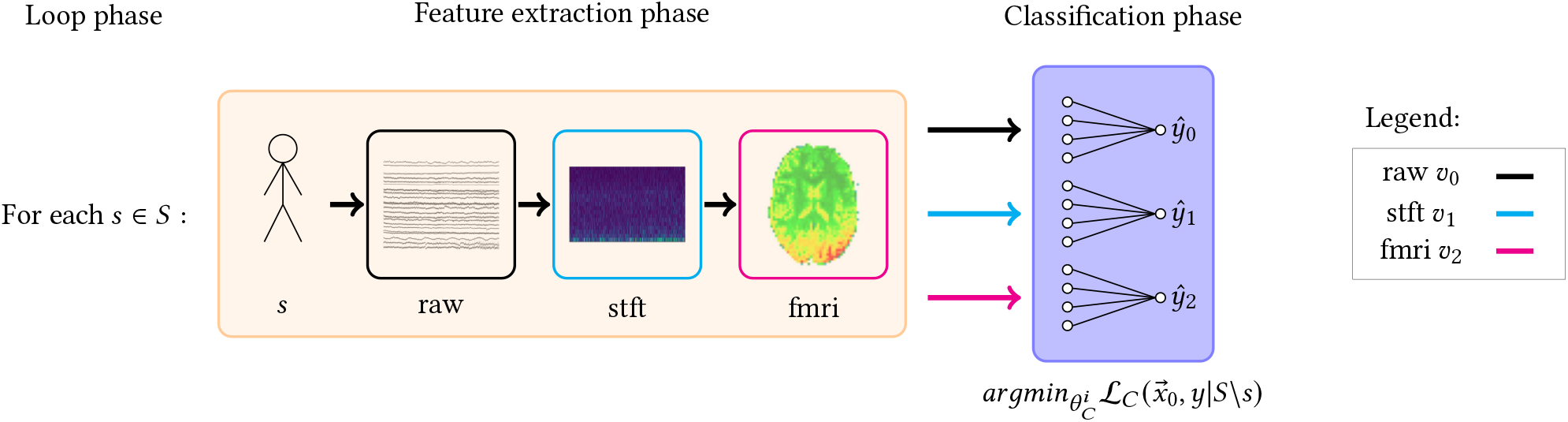
We do a leave-one-individual-out validation, where for each fold we either train a linear classifier with *raw, stft* or *fmri* representations. Each representation has its own validation. The arrows inside the feature extraction phase indicate dependency, that is: an *fmri* representation needs an *stft*; *stft* needs the *raw*; and the *raw*, of course, needs an individual’s recording, denoted with a human figure. For each fold, *s* ∈ *S*, we train a linear classifier without *s*, 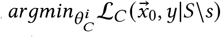. In addition, the *fmri* setting also optimizes 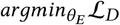. For all individuals/folds the predictions are saved to compute the are under the curve (AUC) against the ground truth.

In terms of synthesis quality, the synthesized fMRI were well defined, meaning that they appeared as fMRI volumes to the human eye. Though, there are ill defined predictions with activity present in the background of the volume. The latter may be due to each model at a fold, converging to different suboptimal parameters.

Consequently, those parameters may lead to different distributions. Nevertheless, the success of the synthesis is encouraging, since it demonstrates the ability of ℒ_*D*_, along with the proposed layer normalization, to maintain the style of the fMRI and at the same time separate the data. The proposed loss also demonstrated to work well in a joint training with an additional classification loss, ℒ_*C*_. We were able to take advantage of a shift invariant function, *cosine*, and process a different EEG dataset by an encoder, that enabled the decoder to project to an fMRI with the learned distribution (the distribution of the NODDI fMRI).

In terms of biclustering, we were able to find biclusters with the parameters for all the resolutions, using the ground truth and the predictions. From the gathered biclusters, we report the best biclusters according to the *lift*. All biclusters have statistical significance, an assurance of the BicPAMS algorithm. In contrast with the classification setting, the biclustering analysis shows us there are patterns with high discriminative power for schizophrenia. Suggesting that models with a better feature engineering would take advantage of these patterns.

## 5 Discussion

Castanho et al. [47] study the application of several biclustering algorithms in fMRI data to uncover statistically significant patterns. One of the algorithms studied was BicPAMS. The biclusters were of the type voxels by time, *D*_2_ × *T*. The authors claim this setup has the leverage of finding patterns that correlate/connect different brain regions over the temporal dimension, a.k.a. functional connectivity. Indeed, it is known that distant brain regions communicate between each other through neuronal pathways. This phenomena can be observed with similar frequencies present in distant EEG electrodes [48]. Note that, we do not consider the temporal dimension of fMRI in our pattern search made by BicPAMS. Thus, we can not make these functional connectivity claims from the synthesized fMRI. Nonetheless, clusters of the form individuals by voxels, *S* × *D*_2_, allow us to assess if there are *constant*^2^ spatial patterns in the fMRI volume that can discriminate schizophrenia. **Outside of brain volume areas, such as background, are not discriminative**. We performed ablation experiments, to ensure no patterns were being found in regions where they were not supposed to exist, such as the background. For this, we collected all these regions of the synthesized volumes, and BicPAMS did not find biclusters with statistical assurances. This experiment rejected the possibility of discriminative information out of brain regions. **Yet, in in brain regions, BicPAMS found several biclusters**, all of them with lift greater than 1.38 for the schizophrenia class. This means, the synthesized fMRIs are able to represent schizophrenia related patterns and do not build patterns in healthy brains. The latter, is particularly encouraging, since healthy brains in this task should not have patterns present.

### The synthesized cerebellum region is present in several biclusters associated with schizophrenia

In the biclusters of the ground truth labels, we found heterogeneous regions in the resolutions. Meaning, finer granularities uncover information that entire lobes, as a whole, do not. This suggests higher resolution volumes may contain relevant patterns that would not be discovered otherwise. To put in perspective, these biclusters in 5 × 5 × 3 volumes were present in the parietal lobe, left temporal lobe and cerebellum; in 10 × 10 × 5 resolution volumes patterns were found in parietal, occipital and left temporal lobes, as well as in the cerebellum region; and in 14 × 14 × 7 biclusters had voxels present in parietal, occipital, frontal and left temporal lobes. Figure 5 illustrates the best biclusters found according to lift. This patterns go in accordance with previous findings reporting that prefrontal and temporal lobes are affected by schizophrenia [49]. On another note, Rahaman et al. [23] made a significant contribution on the application of biclustering in MRI data. They were able to find discriminative MRI patterns for schizophrenia patients. While MRI measures white matter, fMRI records blood supply levels. MRI is not fMRI, but we find it pertinent to relate this study with ours, since biclusters present in regions associated with schizophrenia (gyrus, brainstem) are relevant, independently of the modality. Also, comparing our results, of a synthesized fMRI modality, with an MRI, lets us assess if the synthesis is veracious in a spatial perspective. In their study, a biclustering algorithm was ran on individuals considering nine components (taken from a division of 30 regions of interest using independent component analysis). The biclusters found observed patterns in the gyrus and brainstem parts of the brain. Our different granularity experiments go in accordance with biclusters containing patterns in the cerebellum which is connected to the brainstem. However, we did not report any pattern in the gyrus region. Of course no major claim can be made about the spatial veracity of this inter study correlation, since both our view is synthesized and datasets are different. Additional analyses are needed to check if the activity synthesized in the different regions of the fMRI volume goes in accordance with the dynamics of a real fMRI.

**Figure 5.**
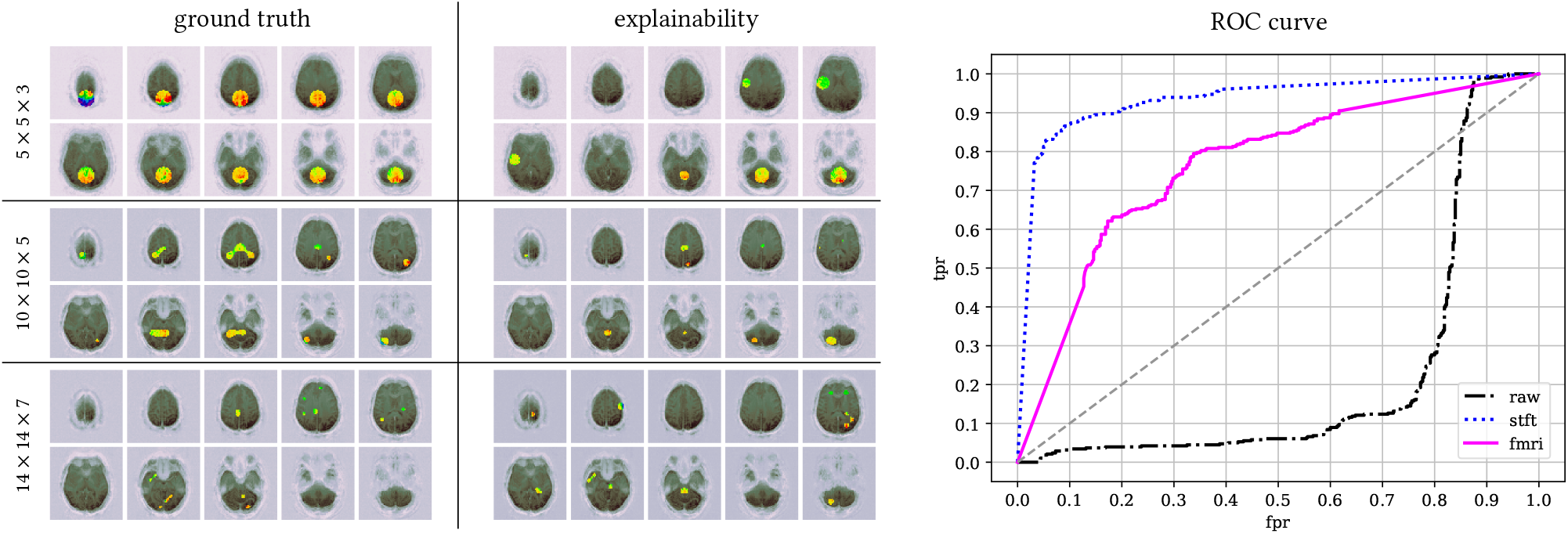
We analyzed resolutions ∈ {5 × 5 × 3, 10 × 10 × 5, 14 × 14 × 7} and gathered the biclusters retrieved for the ground truth and predicted labels. Only the best biclusters (with the best *lift*) are shown in this figure for each setting. On the right side, the receiver operating characteristic (ROC) curve is shown. The *raw* view performed below random, while the *stft* and *fmri* were above. The *stft* is close to a perfect classifier.

In the explainability analysis, we found different biclusters at higher resolutions, but still present in the same regions. In 5 × 5 × 3 resolution volumes, patterns were observed in parietal, occipital, left temporal lobes and cerebellum. At finer granularities, voxels in the frontal lobe were present in the biclusters retrieved for 10×10×5 and 14×14×7 resolutions. Areas uncovered in 5×5×3 were also present at higher resolutions, except for the cerebellum at 14×14×7. These biclusters all reported lifts above 1.2. Lifts in explainability were lower than the ground truth. In general high lift patterns were harder to find in this setting, and as a consequence the *minimum lift* parameter (see Table 1) had to be lowered (relative the ground truth) to relax the search.

### The linear classifier decision making is correlated with the synthesized frontal lobe activity

Doing explainability on a leave-one-individual-out validation schema is difficult. We only have access to the model at the prediction time of the fold. In addition, different folds have different suboptimal parameters. Altogether, these are challenges that we tackled using a pattern mining tool. Pinto et al. [50] used the apriori algorithm to retrieve the explanations of predictions. The idea is that, by finding patterns of data that have statistical assurances for a target, we are explaining predictions. Note however, that the model may not be looking at those patterns to make its decision. What the explainability analysis tells us, is how the synthesized fMRI correlates with the predictions made by the linear classifier. We see them as biclusters that explain the predictions, with statistical assurances. In this setting, BicPAMS found different row sets, because the predictions differ from the ground truth. So, how can we view these biclusters? They discriminate the predictions, a difference seen in the different regions gathered by the biclusters. For instance, explainability biclusters reported the frontal lobe presence, while the ground truth ones did not report this region. Nonetheless, the frontal lobe is associated with problem solving and attention functions, which are recognized as impairments provoked by schizophrenia [21, 22]. This pathology affects the cognitive ability and signatures of the human brain [51]. There is extensive research on the different discriminative patterns that are able to identify this pathology and a lot of research is performed using MRI technologies.

### The synthesized fMRI view is discriminative of schizophrenia

The statistical assurances (lift, support) were higher for the biclusters found in the ground truth analysis. No major comment is made for this observation, as they are different settings, that can not be compared. However, the high discriminative power of the ground truth biclusters for schizophrenia, show that the produced fMRI views have potential for schizophrenia diagnostics. The low AUC, 0.765, of the linear classifier shows us that it is not sufficient for a reliable application of this view. All in all, a linear classifier has no feature engineering properties, but BicPAMS gave us statistical assurances about the discriminative patterns retrieved. Showing us that there is information present in this view, potentially uncoverable by powerful models. We refer this for future work.

Frequency features are highly discriminative of schizophrenia, yet lack interpretability. The linear classifier showed us that it better assessed schizophrenia using the *stft* representation. It is very well known that frequency features are highly discriminative of this pathology [19, 20]. However, we were not able to show the power of the *fmri* view, through this classification setup. Still, this makes sense. The distance from *stft* to the *fmri*, shown in Figure 1, inhibits the gradients of ℒ_*D*_ w.r.t. *θ*_*F*_ to be significant at the top layers. These are the ones close to the *stft* representation. Since the number of epochs is fixed, we hypothesize that not enough time was given in the validation process to show that the *fmri* view is able to perform in comparison to *stft*. We hypothesize that finer regularization strategies, such as *l*_1_-path-norm regularization [52], are needed to allow a faster convergence. Nonetheless, the superiority of *stft* features is due to it being better engineered for schizophrenia. In contrast, the *raw* representation lacks not only in interpretability, but also lacks this engineering property to allow a good performance of the linear classifier. We look at an *fmri* representation and see its potential applicability in a health care setting, since it has higher levels of interpretability and, as previously discussed, highly discriminative patterns. And although, *stft* had a better performance (see ROC curve in Figure 5), it still lacks in the interpretability level. It is essential that a clinical diagnostic be made of simple explanations. Not only has the doctor to understand, but also it is beneficial if the patient fully understands its diagnostic [53]. Explaining a diagnostic based on *stft* features is not tractable for people out of the EEG clinical scope. Not to say that the *fmri* representation is understandable by everyone, but it is easier since it is explained in the spatial (and temporal) domain. EEG is recognized to have low interpretability power [27], however it does not block its use in health care. Our main concern, is projecting this modality to a space where it can better be understood by a human, be it expert or not.

## 6 Conclusion

This study presented a decision making model that is able to produce an *fmri* view from an EEG representation, through the integration of a novel contrastive loss on a state-of-the-art EEG to fMRI synthesis network. The validity of this synthesis task for subsequent diagnosis from single neuroimaging modes/sources is demonstrated for the first in this work, showing delineate predictive accuracy against peer approaches. Our results show that time frequency features, corresponding to an *stft* view, remain as the best option for schizophrenia diagnosis, yet lack interpretability. In turn, we showed, through an exhaustive search framework, that the *fmri* view contains highly discriminative patterns, revealing notable associations of schizophrenia with left temporal, frontal and cerebellum regions. The role of cross-modal augmented neuroimaging views is highlighted as an important subsequent direction.

### Limitations

Note that the synthesized fMRI view analysed in this study has some limitations. We consider the following: Sinusoid-based projections may not be sufficiently expressive to handle all relevant distribution shifts, specially when considering case-control populations; EEG records are mostly suggested for the diagnosis of a limited set of pathologies, for instance, according to our experiments, raw and stft EEG views show poor predictive performance for the classification of Alzheimer’s and Parkinson’s diseases; the synthesized fMRI is only as good as the quality of paired EEG-fMRI records use to train the model. Nowadays, in spite of the emerging set of cohorts with simultaneous EEG and fMRI data, learning limitations related to cohort size and undertaken recording protocols should be carefully noted.

## Data Availability

Data used is available at https://openneuro.org/datasets/ds004000 and https://osf.io/94c5t/.

https://osf.io/94c5t/

## Acknowledgements

This work was supported by national funds through Fundação para a Ciência e Tecnologia (FCT), under the Ph.D. Grant SFRH/BD/5762-/2020 to David Calhas and INESC-ID pluriannual UIDB/50021/2020. We want to thank João Torres for fruitful discussions regarding data imbalance and ways to tackle it. We also give a big thanks to Daniel Gonçalves and Pedro Orvalho for brainstorming along us some ideas of this work. A big thanks to André Patrício for helping us with setting the code in the biclustering analysis.

## Code Availability

Simulation code was written in Python 3. Please refer to the eeg-tofmri Python package and the notebook example (using a subset of the data used) found in this link: classification_contrastive.ipynb.

## Footnotes

1 The kernels of these (downsampling) layers are *k*_1_ ∈ ℝ^7×37×2^ and *k*_2_ ∈ ℝ^7×7×2^. The stride sizes are 3×5×1 and 2×2×1, respectively.

2 Constant patterns are patterns that are equal for every individual.

## Notes

### Competing Interest Statement

The authors have declared no competing interest.

### Funding Statement

This work was supported by national funds through Funda\c{c}\∽ao para a Ci\^encia e Tecnologia (FCT), under the Ph.D. Grant SFRH/BD/5762-/2020 to David Calhas and INESC-ID pluriannual UIDB/50021/2020

### Author Declarations

https://openneuro.org/datasets/ds004000 and https://osf.io/94c5t/

